# The prevalence of low dose aspirin use for prevention of adverse cardiovascular events and its associated factors among patients with diabetes mellitus: A hospital based cross-sectional study

**DOI:** 10.1101/2023.02.13.23285845

**Authors:** Kedir Negesso Tukeni, Ebrahim Umer Mohammed, Nigatu Asfaw Regassa, Eyob Girma Abera

**Affiliations:** Department of Internal Medicine, Jimma University, Jimma, Oromia, Ethiopia; Department of Internal Medicine, Adama Hospital Medical College, Adama, Oromia, Ethiopia; Department of Internal Medicine, Mettu Carl Hospital, Oromia, Ethiopia; Department of Public Health, Jimma University, Jimma, Oromia, Ethiopia; Clinical Trial Unit, Jimma University, Jimma, Oromia, Ethiopia

**Keywords:** low dose Aspirin, Diabetes mellitus, Cardiovascular diseases, primary prevention, secondary prevention

## Abstract

**Background:** Cardiovascular diseases (CVD) are the most common causes of mortality and morbidity among diabetic patients. Aspirin is recommended for primary and secondary prevention of cardiovascular events in patients with diabetics who are eligible for therapy based on active international guidelines. However, these active guidelines are underutilized. Hence, this article helps to assess low dose aspirin use and its associated factors in patients with diabetes mellitus on follow-up at the diabetes clinic of Jimma Medical Center (JMC).

**Method:** A cross-sectional study was conducted among 388 diabetic patients on follow-up at the diabetic clinic of JMC during October 1, 2020 to November 15, 2020. The collected data were cleaned and entered into EpiData version 4.6 then exported to STATA version 16.0 for analysis. Descriptive statistics and multivariable logistic regression was employed to identify the relationship between dependent and independent variables with declaring statistical significance if P value is less than 0.05.

**Result:** Out of the total 388 diabetic patients interviewed, Most of the patients were in the age group of 50-54 (35.8%) with the mean age of 48.8 [48.2, 51.4] years old. About half (48.7%) of them had a history of hypertension. Nearly double of the study participants (47.2%) were utilizing a low dose aspirin. Aspirin was indicated for 80 (20.6%) of the patients despite 21 (26.2%) of them were not using it. It was both indicated and prescribed in 59 (15.2%) of the cases. Older age, longer duration of DM, dyslipidemia, and hypertension were associated with more likely utilization of low dose aspirin.

**Conclusion:** About half of the DM patients were utilizing low dose aspirin, while only one fifth of them were having indications to do so. Furthermore, one fourth of the patients were not taking low dose aspirin for the prevention of cardiovascular events despite they were supposed to use it. Therefore, these findings suggest a greater need for physicians to carefully assess the indications to prescribe aspirin with a clear explanation of the it’s advantages in these specific patient population. Ultimately, future studies should examine the influence of updating guidelines on clinician behaviors to avoid irrational use of aspirin and the association of changing trends in preventive aspirin use with the development of CVD in patients with DM.

## Introduction

The burden of cardiovascular disease (CVD) is substantial in patients with diabetes mellitus (DM), who have a two-to four-fold increase in the incidence of cardiovascular events compared with age- and sex-matched, non-diabetic persons [1]. The prevalence of DM increased from 108 million (4.7%) in 1980 to 425 million (8.5%) in 2017, and it is estimated to be 629 million by 2045 [2]. Morbidity and mortality of CVD in patients with type 2 DM have a risk of death from cardiovascular causes that is two to six times that among persons without DM, and among white Americans the age-adjusted prevalence of coronary heart disease is twice as high among those with type 2 DM as among those without DM [3]. Diabetes-related CVD complications are on the rise that associated with cardiovascular risk factors. Coronary heart disease (CHD) may affect 5% to 8% of type 2 diabetic patients and cardiomyopathy, up to 50% of all patients. Close to 15% of patients with stroke have diabetes, and up to 5% of diabetic patients present with cerebrovascular accidents at diagnosis [4].

In 2015, the Ethiopian Diabetes Association (EDA) and International Diabetic Federation (IDF) reported almost the same number of people living with diabetes in the country 1.33 million and 1.30 million respectively [5]. Various studies were conducted to assess the prevalence of CVD in patients with DM in Ethiopia. The overall prevalence of CVD among T2DM patients was 42.51% in Harari regional state of Ethiopia [6], 15.29% in Jimma Medical Center (JMC) [7]. A systematic review conducted from institutional and community based studies with 5% prevalence of CVD in patients with DM reported [8].

Aspirin has been shown to be effective in reducing cardiovascular morbidity and mortality in high-risk patients with previous myocardial infraction (MI) or stroke (secondary prevention) [9]. In the primary prevention trials, aspirin allocation yielded a 12% proportional reduction in serious vascular events [10]. With its therapeutic dosage of 75–162 mg/day may be considered as a primary prevention strategy in those with Type 1 or Type 2 diabetes who are at increased cardiovascular risk [11]. However, its net benefit in primary prevention among patients with no previous cardiovascular events is more controversial both for patients with diabetes and for patients without diabetes [12]. An association between increases in aspirin dose and risk of adverse events has been confirmed in multiple studies, whereas no such dose relationship has been identified for efficacy. This suggests that following the rapid, acute inhibition of platelet COX-1 with 160 to 325 mg of aspirin, every effort should be made to minimize the long-term dosage [13]. Aspirin has been considered underutilized despite its beneficial effects in primary and secondary prevention of CVD in patients with DM (14). Therefore, this paper assessed the prevalence of low dose aspirin utilization and its associated factors in patients with DM at JMC.

## Methods

### Study design and setting

A cross-sectional study was conducted during October 1, 2020 to November 15, 2020 at the diabetes clinic of JMC, which is located in Jimma town, Southwest Ethiopia, in Oromia region. JMC is a tertiary teaching hospital located in Jimma Town serving a population of over 20 million.

### Source and study population

All Type 1 and Type 2 diabetic patients on follow up at diabetes clinic of JMC from October 1, 2020 - November 15, 2020.

### Inclusion and exclusion criteria

#### Inclusion criteria

All Type 1 and Type 2 diabetic patients on follow up at JMC diabetes clinic.

#### Exclusion criteria

All patients who deny verbal consent to the interview were excluded.

### Sample size determination and sampling technique

The sample size was determined by using single population proportion formula, by considering 50% population proportion of aspirin users, 95% confidence level, 0.05 margin of error and by considering 5% of expected non-response rate, the calculated total sample size was 403. A convenient sampling technique was used taking consecutive patients who come for follow up until the sample size is reached within the study period.

### Data collection instruments and procedure

To assess the prevalence of aspirin use and its associated factors in patients with DM, a structured data collection format was used including the socio-demographic characteristics (age, sex, chart number,marital status, level of education, monthly income, and area of residence) and clinical profiles such as duration of DM, history of CVD and co-administered medications. Prior to data collection, the questionnaire was tested for consistency. The charts of the study population were collected and reviewed after cross-checking chart number on the chart with that on the registration book. Additionally, patients were also interviewed to get the missed data from the charts. Then the data were filled by the investigator into the questionnaire. The data collectors were used personal protective equipment like alcohol-based hand sanitizer, and face mask to reduce the risk of transmission of Corona virus disease (COVID-19) during data collection.

### Operational Definition

**I. Diabetes mellitus**: Refers to a group of common chronic metabolic disorders that share the phenotype of hyperglycemia [14].

**II. Primary prevention**: Initiating appropriate interventions to decrease the likelihood of the first cardiovascular disease occurrence [12].

**III. Secondary prevention**: Initiating the appropriate interventions after a patient suffered an initial cardiovascular insult to minimize the possibility of further insults [12].

**IV. Dyslipidemia**: An abnormal serum concentration and /or abnormal modifications of lipids in the body [12].

**V. Low dose aspirin**: A dose of aspirin in the range of 75- 162 mg [12].

**VI. ASCVD score:** Is a score with different components being used to predict the 10 years’likelihood of developing CVD [12].

**VII. Major Cardiovascular events**: composite of nonfatal stroke, non-fatal myocardial infarction, and cardiovascular death [12].

### Data quality management

All data were collected using a uniform data collection format, and all investigators were trained with the standardized study protocol. The collected data was checked for completeness and consistency on each day of collection. The principal investigator led the overall activities during the data collection period. Questionnaires were prepared in English and back-translated into Afan Oromo/Amharic and translated back into English to check its consistency. The Afan Oromo/Amharic versions were used for data collection after pretesting on 5% of the actual sample size before the data collection. Amendments were made accordingly after pre-testing. Both chart review and patient interview were conducted under strict supervision.

### Data processing and analysis

Data were entered, coded, and cleaned in EpiData manager version 4.6 and then exported to STATA version 16.0 for analysis. Summary findings were presented by tables. Descriptive statistics were used to determine the demographic characteristics and pattern of aspirin utilization. The potential predictor variables were tested in bi-variable logistic regression separately for their association with aspirin utilization. The variables which are significant in the bi-variable analysis at a cut point of P-value of < 0.25 were a candidate for multi-variable logistic regression analysis. Finally, a p-value < 0.05 will be used as a cut-off point for the presence of statistical significance.

### Ethical approval

Ethical clearance was obtained from the Ethical Review Board of Jimma University with a permission obtained from the medical center. Written informed consent from the study participants was obtained and patient confidentiality was ensured during the study period.

## Result

Out of 403 eligible participants, a total of 388 diabetic patients were interviewed for this study as 15 patients were discontinued the interview, yielding a response rate of 96.3%.

### Socio-demographic characteristics

Most of the patients were in the age group of 50-54 (35.8%) with the mean age of 48.8 [48.2, 51.4] years old. The majority of the patients were male (69.1%), and more than half patients were from the rural side of Jimma, Ethiopia (Table 1).

**Table 1:** Socio-demographic characteristics of diabetic patients on follow up at JMC diabetic clinic, Southwest Ethiopia, in 2020.

| Variable | Categories | Frequency (n=388) | Percentage |
| --- | --- | --- | --- |
| Age | <=29 | 55 | 14.2 |
|  | 30-39 | 37 | 9.5 |
|  | 40-49 | 80 | 20.6 |
|  | 50-64 | 139 | 35.8 |
|  | >=65 | 77 | 19.8 |
| Sex | Male | 268 | 69.1 |
|  | Female | 120 | 30.9 |
| Marital status | Married | 331 | 85.3 |
|  | Single | 35 | 9.1 |
|  | Divorced | 10 | 2.5 |
|  | Widowed | 12 | 3.1 |
| Educational status | Never attended school | 111 | 28.6 |
|  | Primary school | 165 | 42.5 |
|  | Secondary school | 58 | 14.9 |
|  | College and above | 54 | 13.9 |
| Residence | Urban | 179 | 46.1 |
|  | Rural | 209 | 53.9 |
| Monthly Income (ETB) | <500 | 17 | 4.4 |
|  | 500-1500 | 61 | 15.7 |
|  | 1501-3000 | 60 | 15.5 |
|  | 3001-5000 | 86 | 22.2 |
|  | >=5001 | 164 | 42.3 |

### Clinical characteristics

Of the total participants, majority of the patients were having 5-9 years history of DM (30.9%), nearly half (48.7%) patients had a history of hypertension. Among the 388 assessed patients, 27 (6.9%) patients were with the history of heartattack, ischemicstroke or peripheral arterial disease (Table 2).

**Table 2:** Clinical characteristics of diabetic patients on follow-up at JMC diabetic clinic, Southwest Ethiopia, in 2020

| Variable | Categories | Frequency | Percentage |
| --- | --- | --- | --- |
| Duration of DM (years) | < =1 | 60 | 15.5 |
|  | 2-4 | 91 | 23.5 |
|  | 5-9 | 120 | 30.9 |
|  | 10-14 | 63 | 16.2 |
|  | >=15 | 54 | 13.9 |
| History of hypertension | Yes | 189 | 48.7 |
|  | No | 199 | 51.3 |
| Current smoker | Yes | 6 | 1.5 |
|  | No | 382 | 98.5 |
| Dyslipidemia | Yes | 110 | 30.1 |
|  | No | 278 | 69.9 |
| CKD*/albuminuria | Yes | 27 | 7.4 |
|  | No | 361 | 92.6 |
| History of heart attack, ischemic stroke or peripheral arterial disease | Yes | 27 | 6.9 |
|  | No | 361 | 93.1 |
| Type of heart disease (n=27) | Myocardial infarction | 7 | 25.9 |
|  | Ischemic stroke | 17 | 62.9 |
|  | Peripheral arterial disease | 3 | 11.2 |

CKD\*, Chronic kidney disease

### Prevalence of low dose aspirin use among diabetics on follow up

Nearly half of patients with DM, 183 (47.2%) were utilizing low dose of aspirin (Fig1).

**Figure 1:**
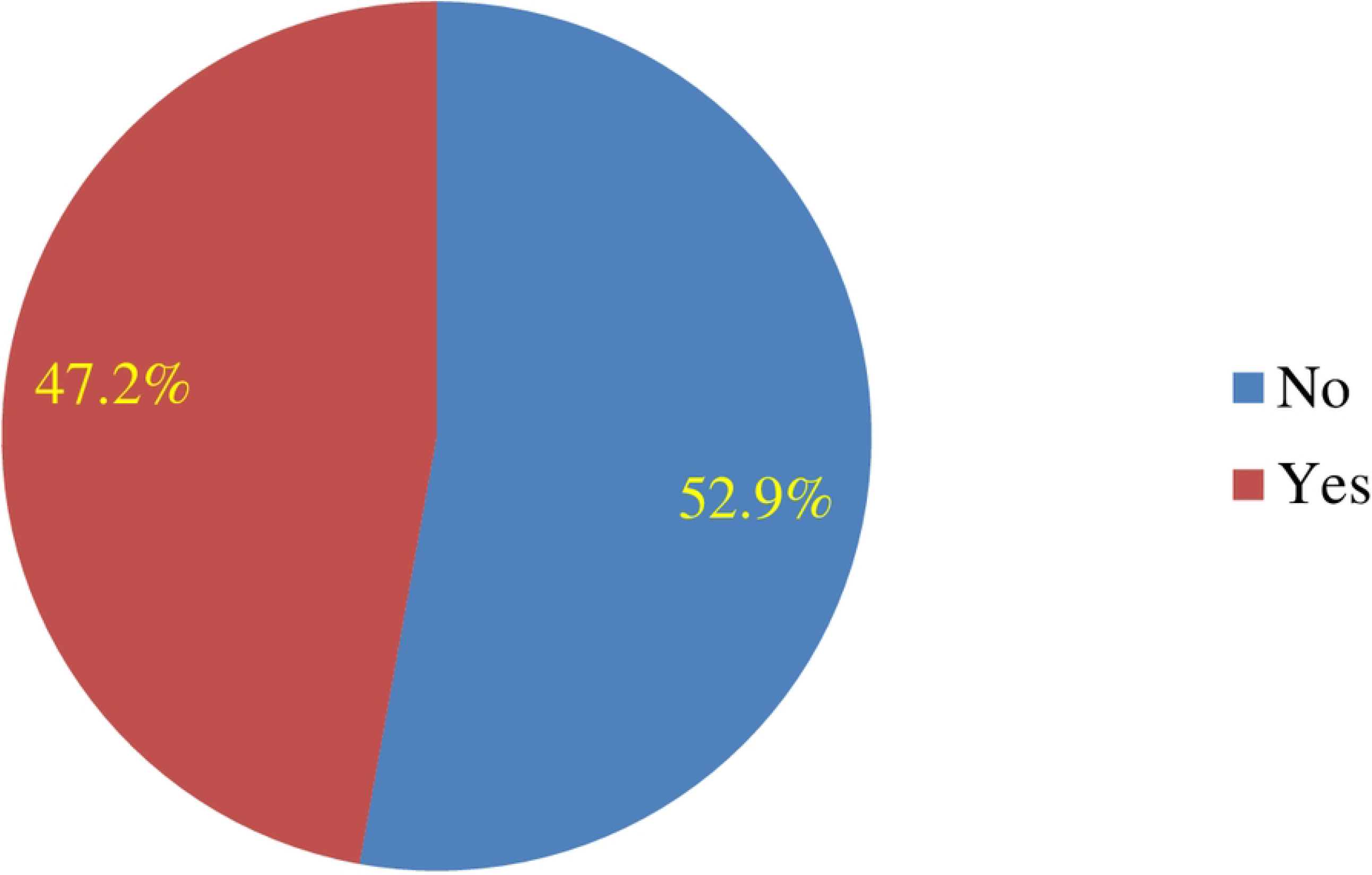
Magnitude of low dose aspirin use among DM patients on follow up.

### Pattern of use of low dose aspirin among diabetics on follow up

The largest proportions (24.2%) of participants taking low dose aspirin were in the age group of 50-64 years old. One hundred thirty-six male participants reported they were taking low dose aspirin. Most of patients (32.2%) with the history of hypertension were using aspirin (Table 3).

**Table 3:** Pattern of use of low dose aspirin use among diabetics on follow up at Jimma Medical Center diabetic clinic, Southwest Ethiopia, in 2020

| Variables | Categories | Have you been taking low dose aspirin |  |
| --- | --- | --- | --- |
|  |  | Yes(%) | No (%) |
| Age group | <=29 | 5 (1.3) | 50 (12.9) |
|  | 30-39 | 4 (1.0) | 33 (8.5) |
|  | 40-49 | 24 (6.2) | 56 (14.4) |
|  | 50-64 | 94 (24.2) | 45 (11.6) |
|  | >=65 | 56 (14.4) | 21 (5.4) |
| Sex | Male | 136 (35.1) | 132 (34.0) |
|  | Female | 47 (12.1) | 73 (18.8) |
| Education | Never attended school | 59 (15.2) | 52 (13.4) |
|  | Primary school | 71 (18.1) | 94 (24.2) |
|  | Secondary school | 23 (5.9) | 35 (9.0) |
|  | College and above | 30 (7.7) | 24 (6.2) |
| Residence | Urban | 96 (24.7) | 83 (21.4) |
|  | Rural | 87 (22.5) | 122 (31.4) |
| Monthly Income (ETB) | <500 | 5 (1.3) | 12 (3.1) |
|  | 500-1500 | 32 (8.3) | 29 (7.5) |
|  | 15001-3000 | 27 (6.9) | 33 (8.5) |
|  | 3001-5000 | 51 (13.1) | 35 (9.0) |
|  | >=5001 | 68 (17.5) | 96 (24.7) |
| Duration of DM (years) | <=1 | 13 (3.4) | 47 (12.1) |
|  | 2-4 | 33 (8.5) | 58 (14.9) |
|  | 5-9 | 72 (18.6) | 48 (12.4) |
|  | 10-14 | 36 (9.3) | 27 (6.9) |
|  | >=15 | 29 (7.5) | 25 (6.4) |
| History of hypertension | Yes | 125 (32.2) | 64 (16.5) |
|  | No | 58 (14.9) | 141 (36.3) |

### Primary and secondary prevention of DM and low dose aspirin use

Among 388 DM patients, 80 (20.6%) of them had an indication to use a low dose aspirin while 53 (13.7%) patients were with a primary indications. However, only 59 (73.8%) of patients were using it, and from those who were using aspirin out of 183 patients, 103 (56.3%) were inappropriately using aspirin. Out of the 27 DM patients who have had an indication to receive low dose aspirin as a secondary prevention, seven (25.9 %) of them were not taking even if they were supposed to use it (Table 4).

**Table 4:** Clients adherence to ADA 2020 guidelines to use low dose aspirin

| Variables | Categories | Taking low dose aspirin |  | Chi square | P-value |
| --- | --- | --- | --- | --- | --- |
|  |  | Yes | No |  |  |
| Primary prevention indicated | Yes | 39 (10.1%) | 14 (3.6%) | 17.194 | 0.000* |
|  | No | 144 (37.1) | 191 (49.2) |  |  |
| Secondary prevention indicated | Yes | 20 (5.2%) | 7 (1.8%) | 8.432 | 0.004* |
|  | No | 163 (42.0%) | 198 (51.0%) |  |  |

*P value*\* <0.05 shows statistical significance

### Reasons for not using low dose aspirin use in indicated cases

Among 80 DM patients with having an indication to use aspirin, 21 (26.2%) clients were not using aspirin, the most (55.2%) commonly cited reason for not using low dose aspirin were being not prescribed (**Fig 2**).

**Figure 2:**
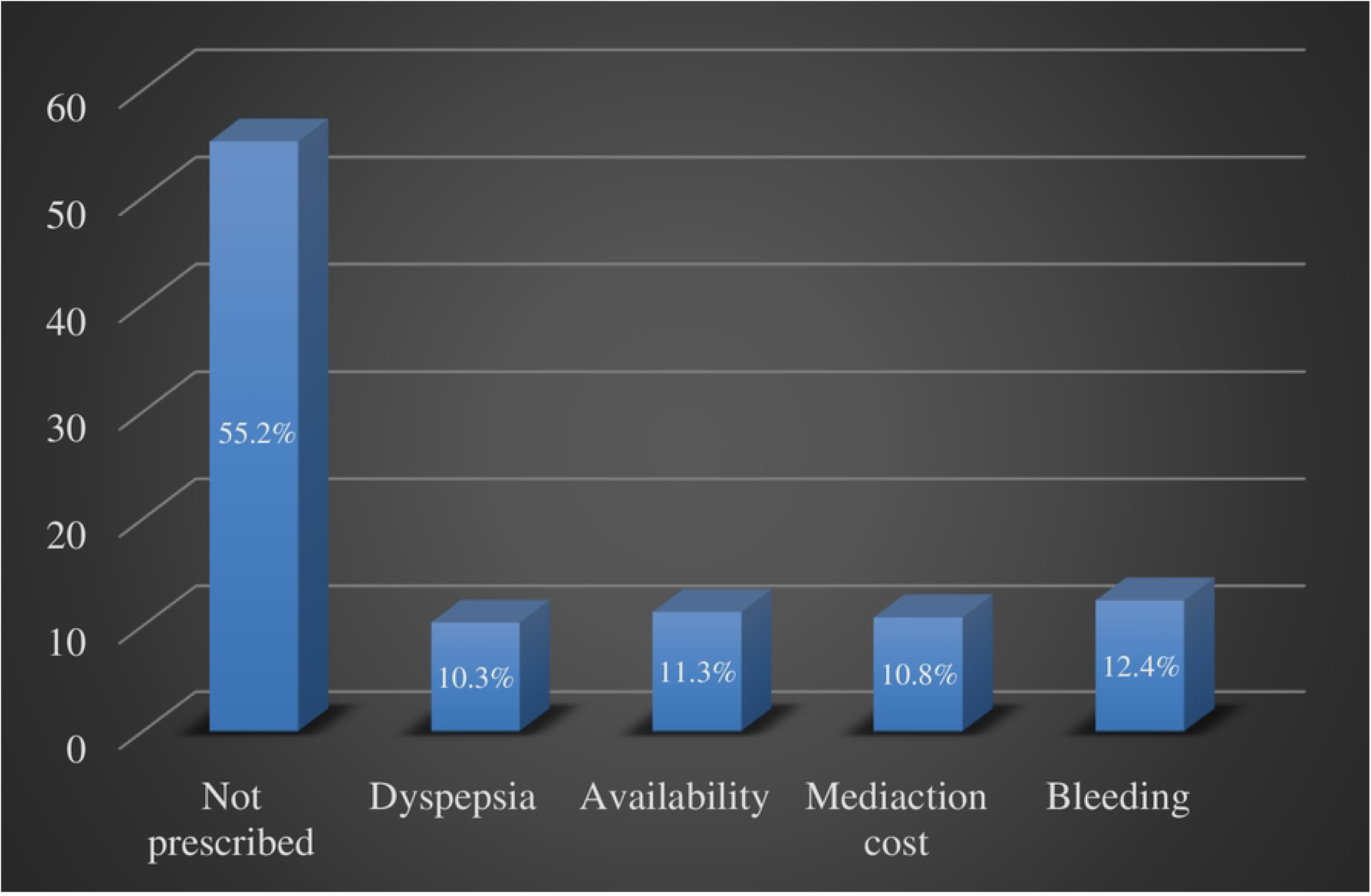
Reason for not using low dose aspirin in indicated case in patients with DM.

### Factors associated with low dose use of aspirin among diabetics

After controlling confounding effects using multivariable logistic regression, factors influencing the use of aspirin among patients with DM were the duration of DM, age group, dyslipidemia, history of hypertension, and chronic kidney disease/albuminuria. Patients who live 10-14 years with DM were six times more likely [AOR 6.25 (95% C.I. 2.33, 16.75)] utilized a low dose aspirin as compared to those living with DM for less than 2 years. In terms of age group, those in the age group of 40-49 years, 50-64 years, and 65 years above were 3.99, 7.28, and 8.51 more likely to utilize a low dose of aspirin as compared to those under 30 years of age respectively. Participants who have a history of hypertension were about twice more likely [AOR 1.97 (95% C.I 1.11, 3.50)] and those who have dyslipidemia were three times more likely [AOR 3.07 (95% C.I 1.60, 5.90)] to utilize a low dose of aspirin compared to their counterparts (Table 5).

**Table 5:** A multi-variable logistic regression model of low dose aspirin use among diabetics on follow up at JMC diabetic clinic, Southwest Ethiopia, in 2020

| Variables | Category | Low dose aspirin |  |  |  |  |  |
| --- | --- | --- | --- | --- | --- | --- | --- |
|  |  | OR | Std. Err. | Z | P>z | [95% C.I] |  |
| Duration of DM (years) | <=1 | 1.00 | (base) |  |  |  |  |
|  | 2-4 | 3.95 | 1.84 | 2.95 | 0.003* | 1.59 | 9.82 |
|  | 5-9 | 5.72 | 3.01 | 5.38 | 0.000* | 3.04 | 8.09 |
|  | 10-14 | 6.25 | 3.14 | 3.64 | 0.000* | 2.33 | 16.75 |
|  | >=15 | 3.49 | 1.76 | 2.49 | 0.013* | 1.30 | 9.35 |
| Monthly Income (ETB) | <500 | 1.00 | (base) |  |  |  |  |
|  | 500-1500 | 2.41 | 1.91 | 1.11 | 0.267 | 0.51 | 11.38 |
|  | 15001-3000 | 2.18 | 1.72 | 0.99 | 0.322 | 0.47 | 10.21 |
|  | 3001-5000 | 3.94 | 2.94 | 1.84 | 0.066 | 0.92 | 16.97 |
|  | >=50001 | 2.01 | 1.47 | 0.95 | 0.341 | 0.48 | 8.40 |
| Age group | <30 | 1.00 | (base) |  |  |  |  |
|  | 30-39 | 1.15 | 0.85 | 0.19 | 0.849 | 0.27 | 4.86 |
|  | 40-49 | 3.99 | 2.29 | 2.42 | 0.016* | 1.30 | 12.28 |
|  | 50-64 | 7.28 | 6.05 | 5.28 | 0.000* | 4.22 | 9.72 |
|  | >64 | 8.51 | 7.65 | 5.14 | 0.000* | 5.86 | 13.85 |
| History of hypertension | Yes | 1.97 | 0.58 | 2.30 | 0.021* | 1.11 | 3.50 |
|  | No | 1.00 | (base) |  |  |  |  |
| Dyslipidemia | Yes | 3.07 | 1.02 | 3.37 | 0.001* | 1.60 | 5.90 |
|  | No | 1.00 | (base) |  |  |  |  |
| CKD<sup>a</sup>/albuminuria | Yes | 1.00 | (base) |  |  |  |  |
|  | No | 1.01 | 0.43 | 0.03 | 0.978 | 0.44 | 2.31 |
| Constant |  | 0.03 | 0.04 | -2.40 | 0.017 | 0.00 | 0.53 |

CKD<sup>a</sup>, chronic kidney disease; *P value*\* < 0.05, statistical significance

## Discussion

This thesis was conducted on 388 diabetic patients to assess the use of low dose aspirin and associated factors for primary and secondary prevention of cardiovascular disease among diabetic patient at JMC from September to November 2020. Of the 388 DM patients, 188 (47.2 %) of them use low dose aspirin. This is a higher rate than the findings in the analysis of the National Health and Nutrition Examination Survey 2011–2012 conducted in the U.S. to examine the use of aspirin for CVD prevention that showed 40.9% reported being told by their physician to take aspirin, with 79.0% (31.6 % of the total) complying [15]. However, a study conducted in US to assess the use of aspirin in diabetes and non-diabetes participants shows a higher use of aspirin utilization among older adults (61.7%) than this study [16]. This is because of the study conducted specifically among older patients with a higher risk rate of indications that possibly increase the utilization of aspirin use. In china, aspirin utilization was 53.5% which is a higher than this study [17].

Among the 27 DM patient with ASCVD, 20 (74.1%) were on low dose aspirin; which is comparable with the study conducted in US to assess recent self-reported regular aspirin use among adults 35 years or older with diabetes in 2001, 74.2 % [18]. From this study, 21.3% of DM patients have had inappropriate aspirin use for primary prevention which is lower than the study conducted in Turkey, 59.2% [19]. However, it was nearly doubled of the inappropriate utilization of aspirin for primary prevention when compared with the study conducted in US which was 11.6% [20]. As per ADA (American Diabetic Association) 2020 guideline [12], among the 388 DM patients, aspirin was indicated for 80 (20.6%) of the patients despite 21 (26.2%) of them were not using it. It was both indicated and prescribed in 59 (15.2%) of the cases. This result is higher when compared to the study conducted in Saudi Arabia, of 312 patients, aspirin was indicated for 17.0% but it was not prescribed; it was both indicated and prescribed in 36.2% of the cases [1]. When compared to the study conducted in India [21], 11% of patients were prescribed with an indication for a primary prevention which is similar with this study, 10.6%; however, 10% of the patients were not utilized despite indicated for a primary prevention which is higher than this study (3.6%) and there was no documentation for the reasons not using aspirin while this study found that 55.2% of patients were reasoned *“not prescribed”*.

In this study, older age, longer duration of DM, dyslipidemia, hypertension, were associated with more likely utilization of low dose aspirin. A similar study done in US, among both in low and high-risk individuals for primary and secondary prevention, higher age groups were more likely to utilize a low dose of aspirin as compared to their counterparts [AOR 5.29 (95% C.I. 3.64, 7.69)] [15] which is similar to this study as age group was found to be a significant predictor of using a low dose of aspirin, which is nearly nine more likely to utilize a low dose of aspirin as compared to those younger age groups [AOR 8.51 (95% C.I. 5.86, 13.85)]. The increase in the prevalence of aspirin use in older populations with diabetes over recent decades may be due to a multitude of factors. Before 2019, aspirin was recommended for older than 50 years without an upper cutoff age limit [22]–[24]. With the updated guideline of the ADA in 2019, aspirin use was discouraged for primary prevention in adults older than 70 years despite aspirin could be considered for patients with high CVD risk and low bleeding risk [25]. In the same year, the American College of Cardiology (ACC) and the American Heart Association (AHA) released updated guidelines that recommended against routine aspirin use for primary prevention in adults older than 70 years [26]. Duration of DM was significantly influencing low dose aspirin utilization. This could be due to detection of risk factors as indications to use low dose aspirin through time during follow-ups. Patients with a history of hypertension were about twice more likely to utilize a low dose of aspirin compared to non-hypertensive DM patients. This is due to its potential risk factor for CVD. As study showed the effect of aspirin in patients with hypertension and diabetes mellitus, there was a 51% reduction in major cardiovascular events in target group ⩽80 mm Hg compared with target group ⩽90 mm Hg (p for trend=0·005) [27].

## Study limitation

The limitation of this study included that patients may not properly remember their age and this may change the recommendations for use of aspirin as the indication for primary prevention of ASCVD is dependent on age. Moreover, we used a cross-sectional study design and cannot draw conclusions related to temporal use of low dose aspirin of any observed associations.

## Conclusion

According to this study, almost half of the DM patients were utilizing low dose aspirin, while only one fifth of them were having indications to do so.And hence more than half of the patients were inappropriately using low dose aspirin. Furthermore, one fourth of the patients were not taking low dose aspirin for the prevention of cardiovascular events despite they were supposed to use it. Therefore, these findings suggest a greater need for physicians to carefully assess the indications to prescribe aspirin with a clear explanation of the advantages of low dose aspirin in these specific patient population. Ultimately, future studies should examine the influence of updating guidelines on clinician behaviors to avoid irrational use of aspirin and the association of changing trends in preventive aspirin use with the development of CVD in patients with DM.

## Data Availability

All relevant data are within the paper.

## Acknowledgement

We would like to express our sincere and deep-rooted thanks to study participants and data collectors for the support provided since the training upon the final submission of the manuscript.

## Conflict of interest

The authors report no conflicts of interest in this work.

## Funding

This research received no specific grant from any funding agency in the public, commercial, or not-for-profit sectors.

## Authors’ contribution

Conceptualization: KNT,NAR; Writing – original draft: KNT,NAR; Statistical analysis: EG; Data Curation: NAR, EUM; Writing – review & editing: KNT, EGA

## Notes

### Competing Interest Statement

The authors have declared no competing interest.

### Author Declarations

The IRB of Institute of Health has reviewed the research project on Assessment of low dose ASA use for prevention of CVD and associated factors among diabetic out-patients at Jimma Medical Center, and notified that it meets Ethical and Scientific standards outlined in the National and International guidelines. Hence, we are pleased to inform you that your research protocol is ethically cleared. Ethical clearance was obtained from the Ethical Review Board of Jimma University with a permission obtained from the medical center.

